# Implementation and Adherence to Regular Asymptomatic Testing in a COVID-19 Vaccine Trial

**DOI:** 10.1101/2024.02.28.24303505

**Authors:** Lucy R Williams, Katherine R W Emary, Daniel J Phillips, Jodie Hay, Jessica P J Larwood, Maheshi N Ramasamy, Andrew J Pollard, Nicholas C Grassly, Merryn Voysey

## Abstract

**Background:** For pathogens which cause infections that present asymptomatically, evaluating vaccine efficacy (VE) against asymptomatic infection is important for understanding a vaccine’s potential epidemiological impact. Regular testing for subclinical infections is a potentially valuable strategy but its success hinges on participant adherence and minimising false positives. This paper describes the implementation and adherence to weekly testing in a COVID-19 vaccine trial.

**Methods:** COV002 was a phase 2/3 trial assessing the ChAdOx1 nCoV-19 vaccine against SARS-CoV-2. Asymptomatic infections were detected using weekly self-administered swabs for RT-PCR testing. We analysed adherence using mixed-effects regression models and estimated the probability of true and false positive asymptomatic infections using estimates of adherence and testing characteristics.

**Findings:** 356,551 tests were self-administered by 10,811 participants during the 13-month follow-up. Median adherence was 75.0% (IQR 42·6-90·9), which translated to a 74·5% (IQR 50·9-78·8) probability of detecting a positive asymptomatic infection during the swabbing period, and between 21 and 96 false positives during VE evaluation. The odds of returning a swab declined by 8% per week and further after testing positive and unblinding. Adherence was higher in older age groups, females and non-healthcare workers.

**Interpretation:** The COV002 trial demonstrated the feasibility of running a long-term regular asymptomatic testing strategy. This information could be valuable for designing future phase III vaccine trials in which infection is an outcome.

**Funding:** UK Research and Innovation, National Institutes for Health Research (NIHR), Coalition for Epidemic Preparedness Innovations, NIHR Oxford Biomedical Research Centre, Thames Valley and South Midland’s NIHR Clinical Research Network, AstraZeneca.

**Research in context:** *Evidence before this study:* Regular testing for asymptomatic infections in clinical trials is useful for evaluating the role of candidate vaccines or drugs in preventing infection. While there is extensive research on loss to follow-up in clinical trials, there is minimal research on adherence to repeat clinical trial procedures. During the COVID-19 pandemic, regular asymptomatic testing was used for surveillance and contact tracing in isolated populations, and in two SARS-CoV-2 vaccine trials. We searched PubMed from database inception to Dec 17, 2023 using the following search terms (title or abstract) for articles published in English: (Adherence OR Compliance OR Uptake OR Implementation) AND (Repeat test* OR regular test* OR weekly test* OR monthly test* OR serial test*) AND (“covid*” OR “COVID-19*” OR “SARS-CoV-2*” OR “Severe Acute Respiratory Syndrome Coronavirus 2”), and reviewed the identified publications. Identified studies demonstrated the feasibility over short time periods of regular asymptomatic testing in hospital, care home, university, school and workplace settings. A small number evaluated differences in adherence by socio-demographic characteristics, mainly highlighting increased age as a predictor of adherence. No studies evaluated adherence in clinical trial settings or predictors of adherence over time.

*Added value of this study:* We evaluated the feasibility and adherence to regular asymptomatic testing in a phase III trial of the ChAdOx1 nCoV-19 vaccine against SARS-CoV-2. We demonstrated high adherence across trial participants over a year of follow-up, but significant differences across socio-demographic characteristics. Adherence was highest in older age groups, females and non-healthcare workers, and declined most strongly over time in younger age groups. We show how the frequency of testing can be translated to an estimated probability of a false positive and negative infections.

*Implications of all the available evidence:* The overall evidence suggests that regular asymptomatic testing is a feasible strategy for tracking the risk of infection for diseases with a high proportion of subclinical infections. Strategies to support subpopulations to maintain adherence over prolonged periods of time may be necessary, and consideration needs to be given to the optimal time over which this type of intensive sampling provides valuable data. Further research into the effect of variation in adherence to regular testing on vaccine efficacy estimates would be valuable.

## Introduction

Many pathogens cause infections that present asymptomatically in a proportion of infected persons, such as typhoid, malaria, dengue, and SARS-CoV-2.^1-3^ For these pathogens, understanding whether candidate vaccines prevent initial infection, progression to symptomatic illness, or some degree of both, is important for deciphering their potential epidemiological impact.^4^ However, measuring vaccine efficacy (VE) against asymptomatic or overall infection can be challenging when traditional methods of screening symptomatic trial participants will not detect a proportion of infections.

A common approach for measuring asymptomatic infection is serological testing at predefined intervals during and at the conclusion of a trial.^5,6^ However, this is only possible if natural and vaccine derived immune responses can be distinguished, and it relies on asymptomatic infections inducing a detectable, long-lasting antibody response. Alternatively, asymptomatic infections can be detected in real-time with regular nucleic acid or antigen testing during the trial.^4^ This can provide additional insights into pathogen load, duration of infection and offers the potential for sequencing isolates.^7^ However, regular testing is resource-intensive and logistically challenging, so it is not usually employed in phase III vaccine trials.

The COVID-19 pandemic stimulated extensive research in the search for a safe and efficacious vaccine. While the primary goal of the phase III vaccine trials was to identify a vaccine that prevented symptomatic and severe disease, a vaccine that prevented asymptomatic infection and onwards transmission would have a much greater public health benefit.^8^ Numerous SARS-CoV-2 vaccine trials employed serological strategies for the measurement of VE against asymptomatic infections.^6,9-11^ In these trials, a subset of trial participants had serological tests for anti-nucleocapsid antibodies at pre-specified time intervals. This allowed for seroconversion after infection to be identified for trials of vaccines based on the spike protein. In two trials, asymptomatic infections were detected using regular PCR testing of asymptomatic participants.^7,12^ These were the Bharat Biotech-sponsored trial of BBV152, where participants were asked to test monthly, and the University of Oxford and AstraZeneca-sponsored trial UK trial, COV002, of ChAdOx1 nCoV-19, where testing was performed weekly.

The feasibility of these testing strategies in large phase III trials, particularly in a pandemic setting, was not known prior to their implementation. Given the high commitment required for these protocol procedures, adherence was expected to be incomplete and decline over time.^13^ Low adherence for asymptomatic testing could introduce a bias in VE estimation.^14^ This is possible when a vaccine prevents symptom development, because the action of the vaccine in preventing symptoms also reduces the probability of that infection being detected. This means that VE against overall infection can be overestimated. Conversely, if frequent testing is conducted with high adherence, false positives become more likely, which can introduce an opposing bias.^14,15^

This paper aims to describe the implementation of regular asymptomatic testing in the COV002 trial and to evaluate adherence to testing over time. Additionally, we examine factors associated with lower test return rates and estimate the probabilities of detecting true and false positive asymptomatic infections. By investigating these aspects, we can gain valuable insights into the challenges and opportunities associated with a regular testing strategy to better inform future trials.

## Methods

### Study Design

COV002 was a single-blind, randomised controlled phase 2/3 trial assessing the efficacy of the intramuscular ChAdOx1 nCoV-19 vaccine using a licensed intramuscular MenACWY vaccine as a comparator. Version 14.0 of the protocol was published as a supplementary appendix to the interim efficacy analysis, which combined data from the four randomised controlled trials testing ChAdOx1 nCoV-19 efficacy.^16^ Briefly, 10,811 adults aged ≥18 years were recruited and vaccinated at 19 sites across England, Wales, and Scotland between 31^st^ May and 24^th^ November 2020. Recruitment of individuals with high occupational exposure risk to SARS-CoV-2 were prioritised.

Participants were enrolled into several study groups for the phase 2 (immunogenicity) and 3 (efficacy) components separately. Other subgroups enrolled included participants who had received a previous ChAdOx1 viral-vectored vaccine (from non-Covid trials) and individuals who were HIV positive. Participants were randomised to receive either the ChAdOx1 nCoV-19 vaccine or the licensed MenACWY vaccine according to the specific ratio for their allocated group.

### Unblinding procedures

The UK COVID-19 vaccination programme started on 8^th^ December 2020 according to a priority schedule related to age, vulnerability and occupational risk. Participants were asked to remain blinded as to their trial vaccine allocation until the day of their NHS COVID-19 vaccination at which point they were unblinded.

### Weekly self-swabbing

All COV002 participants were asked to perform a home test once per week starting one week after their prime vaccination. The Department of Health and Social Care provided home swab kits to the COV002 trial which were distributed directly to participants at study visits by sites. Participants were given standardised instructions and asked to swab their nose and throat. Initially they were asked to do this for 8 weeks, but as feasibility and security of kit supply was established, the duration of self-swabbing was extended until their withdrawal or 30^th^ June 2021, whichever came sooner for each individual. Details of the test packaging, processing, and matching to trial participants are provided in the Supplementary Methods.

### Analysis populations

This analysis includes all weekly swabs returned by all enrolled participants between enrolment and the earliest of 30^th^ June 2021 and withdrawal. This incorporates all swabs regardless of test outcome and development of symptoms. We also defined the “VE analysis population”, which included all enrolled participants who were enrolled in efficacy rather than immunogenicity cohorts who were eligible for VE analysis, during their period of VE evaluation, from two weeks post second dose to the first of infection, unblinding, withdrawal, or 30^th^ June 2021. In this subset, participants were ineligible if they i) were not seronegative at baseline, ii) did not receive two doses, iii) were censored before the start of follow-up (unblinded or received a positive swab).

### Statistical analyses

Each participant’s follow-up was divided into weeks, and one test was expected to be returned each week, from one week post enrolment. An expected test was recorded as provided if it was given between day 0 and 7 of that week, or if two tests had been taken in the week prior or before, the test was recorded as having been taken in the week without a recorded test.

Multivariable logistic mixed regression models were performed to evaluate predictors of adherence to regular PCR testing. The outcome was the presence of an expected weekly asymptomatic swab. Predictor were selected from a set of candidate variables: age (<25 years, 25-34 years, 35-44 years, 45-54 years, 55-64 years, ≥65 years), sex, occupation (healthcare-worker [HCW] seeing ≥1 COVID-19 patient, HCW not seeing COVID-19 patients, essential non-HCW, non-essential non-HCW), ethnicity (white and non-white), BMI category (underweight, healthy weight, overweight, obese), comorbidities (yes/no), inclusion in VE analysis (yes/no), number of weekly UK COVID-19 cases, time since first dose (weeks), and binary variables for whether the swab was expected before the start of VE follow-up (two weeks post second dose), post-unblinding, post-positive swab, and post-December 8^th^, when the UK vaccine rollout began. All variables and their interactions with time were added to a full multivariable model, except the binary time-dependent variables which were added without interaction. The final variables were selected by backwards selection, to a threshold p-value of 0·05. A random effect on the intercept was added for each participant, and an autoregressive covariance matrix was used to account for correlation over time. We present the results of the full multivariable model with all interactions with time, as well as a multivariable model with only additive predictors, to facilitate interpretation of each variable’s effect over the duration of follow-up.

The probability of asymptomatic infection detection with weekly PCR swabbing for different levels of adherence was estimated using data on PCR test sensitivity over time since infection^17^ (Supplementary Methods, Supplementary Table 1-2). The individual-level probability of receiving a false positive was estimated using the number of asymptomatic tests returned and the test specificity.

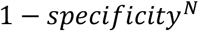

where N represents the number of tests. Test specificity was based on published estimates for a low prevalence setting.^18^ A central estimate of 99·945% was used as a best approximation, and upper and lower bounds of 99·99% and 99·91% were tested to evaluate sensitivity to the assumed specificity. We estimated the number of false positives through the trial by summing over the individual probabilities of a false positive.

## Results

Almost all enrolled participants (10462, 96·8%) returned at least one asymptomatic swab between their first dose and the end of the 57-week swabbing period (median = 27 weeks). 8548 participants were eligible for the VE subset, and 8156 (95·4%) of these provided at least one asymptomatic swab during the period of VE follow-up. The characteristics of all enrolled participants are given in Table 1, and the VE subset in Supplementary Table 3. A total of 356,551 weekly swabs were returned, with 107,641 collected by eligible participants during the period of VE evaluation. 97·7% of tests were negative (Supplementary Table 4).

**Table 1.** Health and socio-demographic characteristics of all participants enrolled in the COV002 phase III vaccine trial.

|  | All enrolled participants (N=10811) | Low adherence participants: <75% swabs returned (N =5383) | High adherence participants: ≥75% swabs returned (N = 5428) |
| --- | --- | --- | --- |
| **Age, years** |  |  |  |
| <25 | 585 (5·4) | 482 (9·0) | 103 (1·9) |
| 25-34 | 2396 (22·2) | 1687 (31·3) | 709 (13·1) |
| 35-44 | 2557 (23·7) | 1508 (28·0) | 1049 (19·3) |
| 45-54 | 2596 (24·0) | 1177 (21·9) | 1419 (26·1) |
| 55-64 | 1141 (10·6) | 337 (6·3) | 804 (14·8) |
| ≥65 | 1536 (14·2) | 192 (3·6) | 1344 (24·8) |
| **Sex** |  |  |  |
| Male | 4369 (40·4) | 2072 (38·5) | 2297 (42·3) |
| Female | 6442 (59·6) | 3311 (61·5) | 3131 (57·7) |
| **Ethnicity** |  |  |  |
| White | 9967 (92·2) | 4818 (89·5) | 5149 (94·9) |
| Asian | 552 (5·1) | 370 (6·9) | 182 (3·4) |
| Black | 49 (0·5) | 30 (0·6) | 19 (0·4) |
| Other | 243 (2·2) | 165 (3·1) |  |
| **BMI kg/m<sup>2</sup>** |  |  |  |
| Underweight (<18·5) | 106 (1·0) | 42 (0·8) | 64 (1·2) |
| Healthy weight (18·5 to 24·9) | 4762 (44·0) | 2306 (42·8) | 2455 (45·2) |
| Overweight (25 to 29·9) | 3773 (34·9) | 1821 (33·8) | 1954 (36·0) |
| Obese (≥30) | 2170 (20·1) | 1214 (22·6) | 955 (17·6) |
| **Occupation<sup>1</sup>** |  |  |  |
| Non-essential | 1693 (15·7) | 329 (6·1) | 1364 (25·1) |
| Essential, non-HCW | 2121 (19·6) | 937 (17·4) | 1184 (21·8) |
| HCW (0 COVID-19 patients) | 4805 (44·4) | 2674 (49·7) | 2131 (39·3) |
| HCW (1+ COVID-19 patients) | 2192 (20·3) | 1443 (26·8) | 749 (13·8) |

Data are N (%). BMI = Body mass index, HCW = Healthcare worker. <sup>1</sup>Non-essential occupation classified as: Unemployed, Retired, Student, Hospitality/retail workers in non-essential shops, Construction workers and labourers, or Other occupation. Essential, non-HCW occupation = Cleaning and domestic workers, Drivers and transport workers, Public safety workers (e.g. police, fire services, security), Religious workers, and Retail workers in essential shops (e.g. food, chemists, banks).

The median adherence to weekly PCR testing was high (75·0%, interquartile rage [IQR] 42·6-90·9%), with 43·4% of participants returning at least 80% of expected swabs (Figure 1). Adherence was particularly high during the VE follow-up, where 63·9% of eligible participants returned at least 80% of expected swabs (Supplementary Figure 1). There was a gradual and non-linear decline in the proportion of expected swabs returned over calendar time (Figure 2) and time since prime vaccination (Supplementary Figure 2). In the first week after prime vaccination, 84·2% of participants returned their expected swab. Adherence maintained above 80% for 7 weeks, 60% for 33 weeks, and 40% for 49 weeks. The rate of decline in adherence reduced in Autumn 2020, but there was a notable drop in Winter 2020, coinciding with a number of non-pharmaceutical interventions and the beginning of UK vaccine rollout.

**Figure 1.**
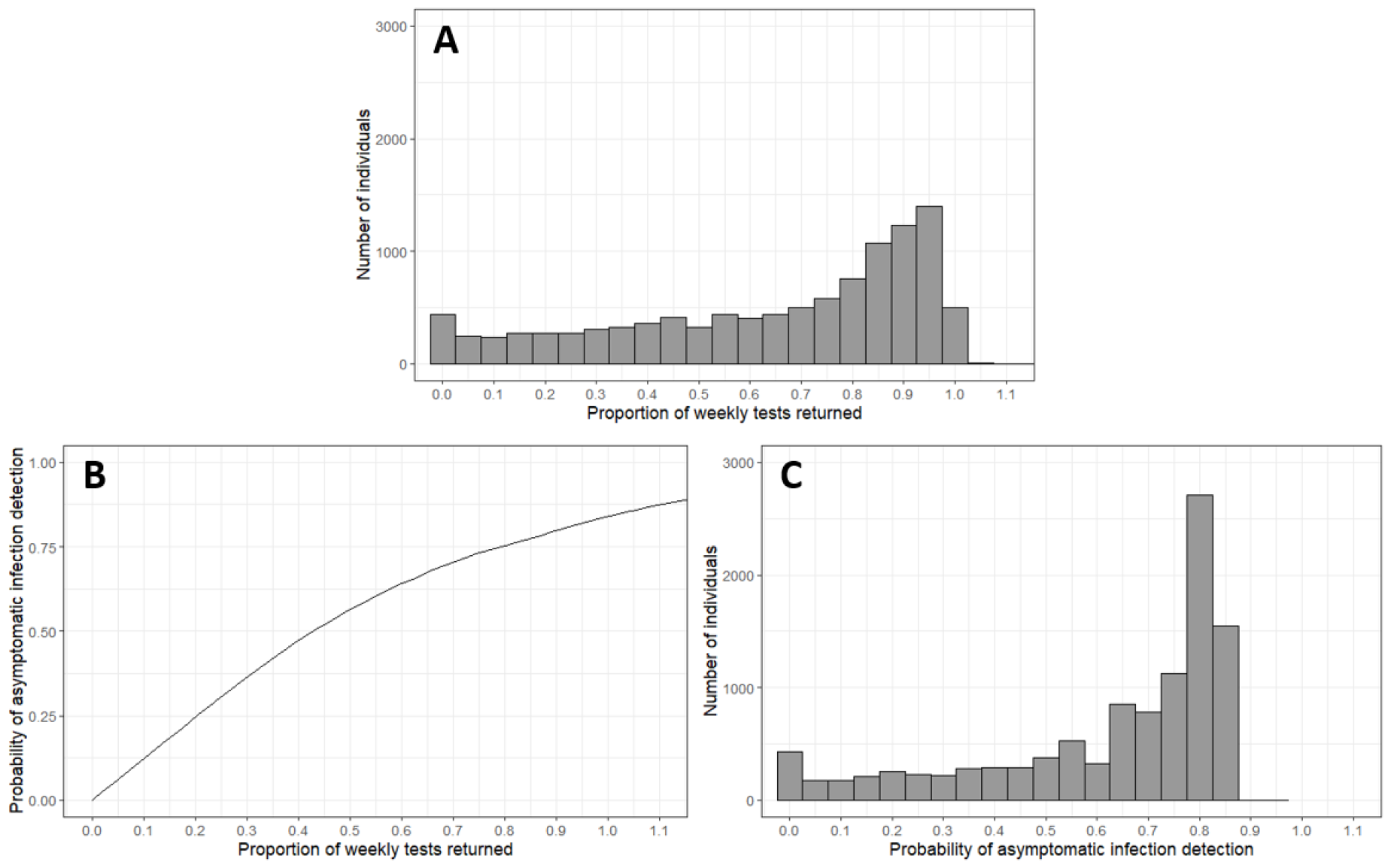
Adherence and estimated probability of asymptomatic infection detection in all enrolled participants in the COV002 trial. A) Histogram of participants’ adherence to weekly testing, B) Assumed relationship between proportion of weekly tests returned and the probability of asymptomatic infection detection, C) Histogram of the participants’ estimated probability of asymptomatic infection detection.

**Figure 2.**
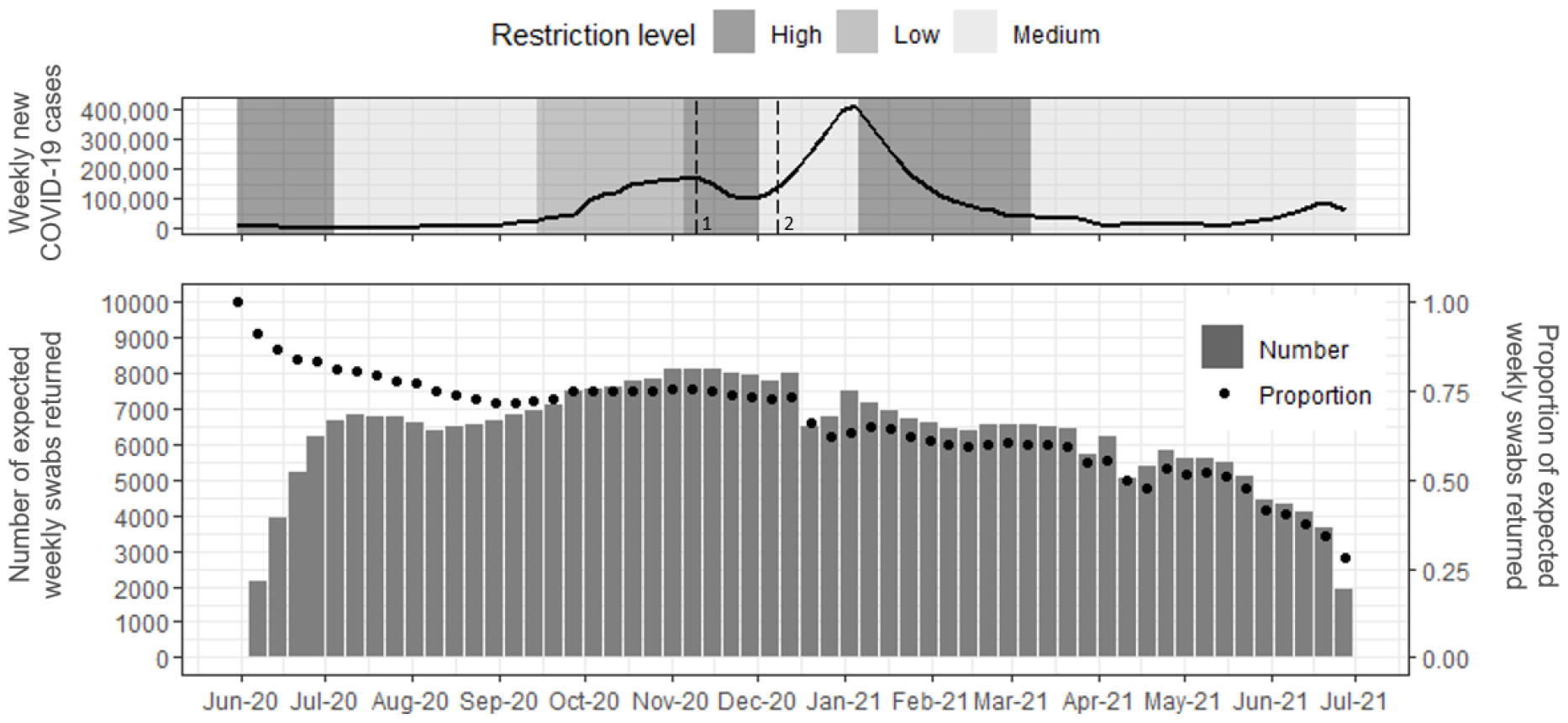
Number and proportion of swabs returned over calendar time, compared to UK epidemic trajectory (weekly number of reported COVID-19 cases) and restrictions. Dotted lines represent the release of the first vaccine results (1) and the beginning of the UK vaccine rollout, when trial participants were also notified that they would be unblinded (2). Stages of restriction levels, in chronological order: First national lockdown (March to June 2020), Minimal lockdown restrictions (July to September 2020), Tiered restrictions (September to October 2020), Second national lockdown (November 2020), Tiered restrictions (December 2020), Third national lockdown (January to March 2021), Phased exit from lockdown (March to July 2021).

We estimated an 8·1% reduction in the odds of swab return (OR 0·919, 95% CI 0·917 to 0·921) per week since first dose (Figure 3). Adherence also varied by a number of socio-demographic characteristics. Age was the strongest predictor, with older age groups significantly more likely to return a weekly swab than younger age groups (OR 65+ years vs <25 years = 25·6, 95% CI 20·0 to 32·9). HCWs, particularly those seeing COVID-19 patients were less likely to return a swab than non-HCWs, while people with comorbidities were slightly more likely to adhere to swabbing (OR comorbidity vs no comorbidity = 1·1, 95% CI 1·0 to 1·3). Controlling for time, participants were more likely to return a swab while VE was being evaluated. After unblinding, the odds of returning a swab were 32% lower than during VE follow-up, and after receipt of a positive swab, 56% less likely. We could not evaluate the association between the weekly number of COVID-19 cases and adherence because the week since prime and number of cases variables were both confounded by calendar time. There was no statistically significant difference in adherence between essential and non-essential non-HCWs.

**Figure 3.**
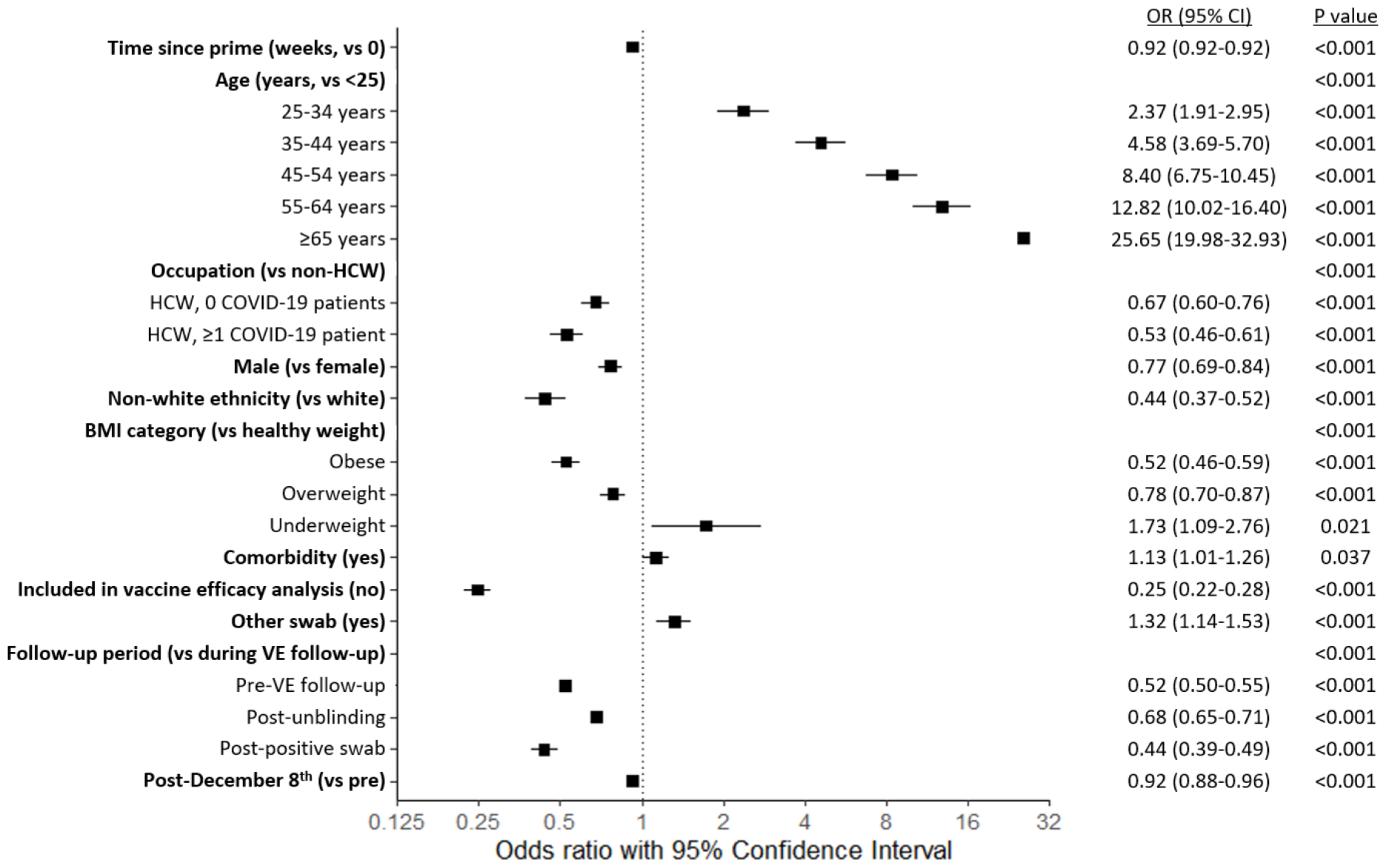
Forest plot of predictors of adherence to regular testing in the COV002 trial. Variables used in multivariable model: Time since prime (weeks), Age (years, <25, 25-34, 35-44, 45-54, 55-64, ≥65), Occupation (non-healthcare worker, healthcare worker not seeing COVID-19 patients, healthcare worker seeing one or more COVID-19 patients), Sex, Ethnicity (white, non-white), BMI (underweight, healthy weight, overweight, obese), Comorbidity, Inclusion in vaccine efficacy (VE) analysis, Other non-weekly swab taken that week, Follow-up period (pre-vaccine efficacy follow-up, during vaccine-efficacy follow-up, post-unblinding, post-positive swab), and pre/post December 8^th^. Reference characteristics: 1 week since prime, <25 years, non-essential worker, Female, white ethnicity, healthy weight, no comorbidities, participant enrolled in efficacy cohort, participant included in VE analyses, no other swabs taken that week (e.g., symptomatic swab/workplace swab), swab expected during VE follow-up, pre-December 8th (start of UK vaccine rollout). Participants were included in VE analysis if they were enrolled in the efficacy cohort, were seronegative at baseline and did not test positive before the start of VE follow-up. Group P values calculated with type III ANOVA. HCW = Healthcare worker, BMI = Body mass index, VE = vaccine efficacy.

The change in adherence over time also varied across socio-demographic groups (Figure 4, Supplementary Table 5). The decline over time was greatest in the youngest age groups (Figure 4A), with the odds of returning a swab 99% lower 40 weeks after primary vaccination, compared to 1 week post first dose (OR week 40 vs week 1 = 0·010, 95% CI 0·007 to 0·012). Although the decline was also significant in the oldest age group of participants 65 years and above, it was more moderate with a 78% reduction between week 1 and week 40 after the first dose (OR week 1 vs week 40 = 0·218, 95% CI 0·173 to 0·275). Notable declines in adherence over time were also seen for BMI category and inclusion in VE analysis. Model predicted probabilities of returning an expected asymptomatic test over time are shown in Supplementary Figure 3.

**Figure 4.**
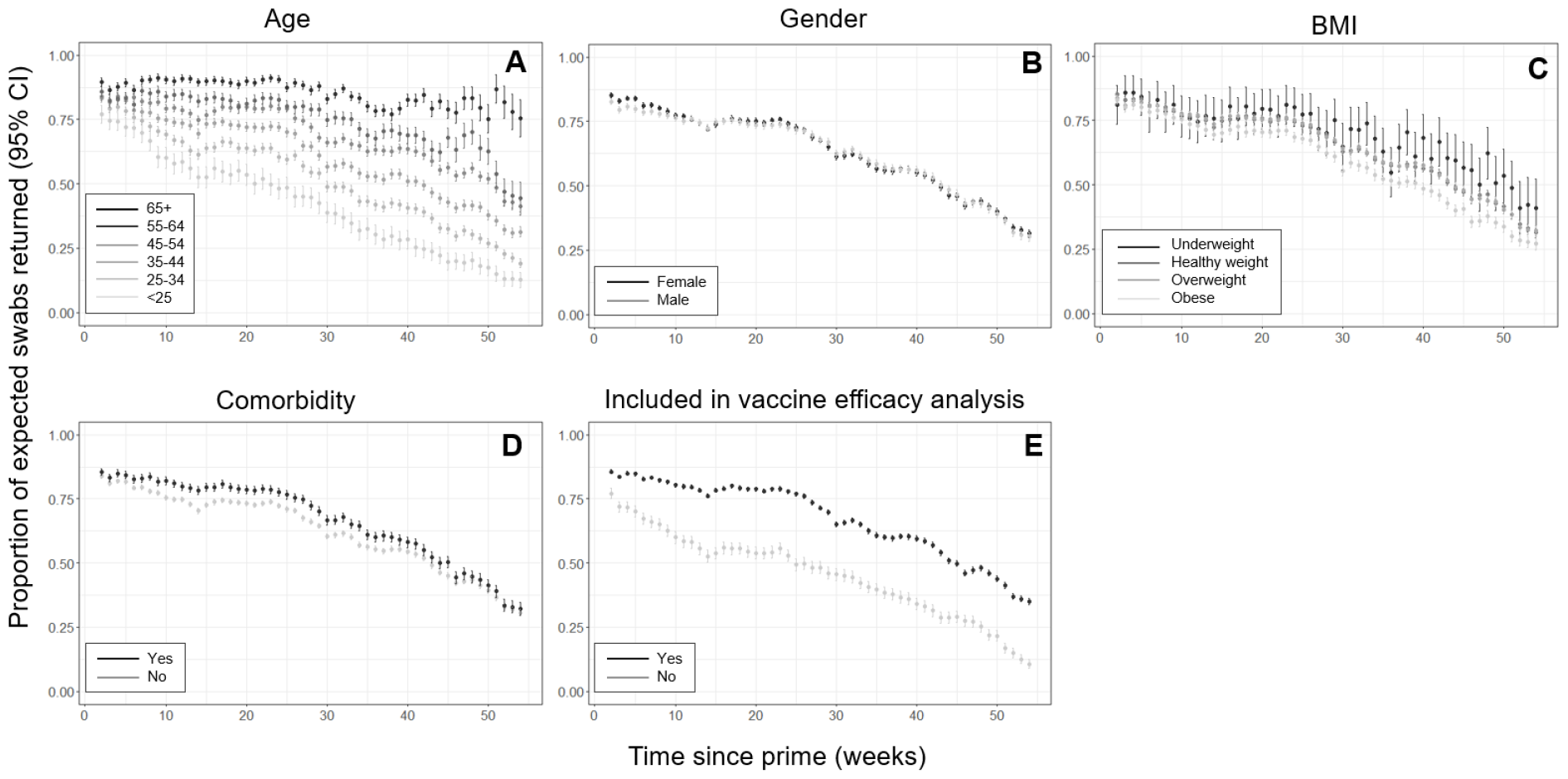
Observed proportion returning an expected asymptomatic test over time, by different socio-demographic variables. A) age, B) gender, C) BMI, D) Comorbidity, E) Inclusion in vaccine efficacy (VE) analysis. Participants were included in VE analysis if they were enrolled in the efficacy cohort, were seronegative at baseline and did not test positive before the start of VE follow-up.

The average probability of asymptomatic infection detection with weekly swabbing if all expected swabs are returned is 83% (Figure 1B). If an individual completed three quarters of their swabs, their probability of detection was estimated at 74%, while a 50% return rate was estimated at a 56% probability. Accounting for test sensitivity and incomplete adherence to testing, we estimated the median probability of asymptomatic infection detection over time across all participants enrolled at 74·5% (IQR 50·9-78·8, Figure 1C). Across participants eligible for VE analysis during the period of VE follow-up, the estimated probability was 78·8% (IQR 70·3-83·4, Supplementary Figure 1B).

The estimated number of false positives was sensitive to the assumed test specificity (Figure 5). The probability of receiving a false positive test result for a participant returning the median number of 36 tests was estimated at 3·19% if using the lower bound for test specificity (99·91%), and 0·72% if using the upper bound (99·98%). Assuming the central test specificity (99·945%), the estimated number of false positives in all enrolled participants was 194, but this ranged between 71 and 315 for the upper and lower bounds for the test specificity. In the VE subset for the period of VE analysis, we estimate that between 21 and 96 false positives were detected, with a central estimate of 59 (Supplementary Figure 4).

**Figure 5.**
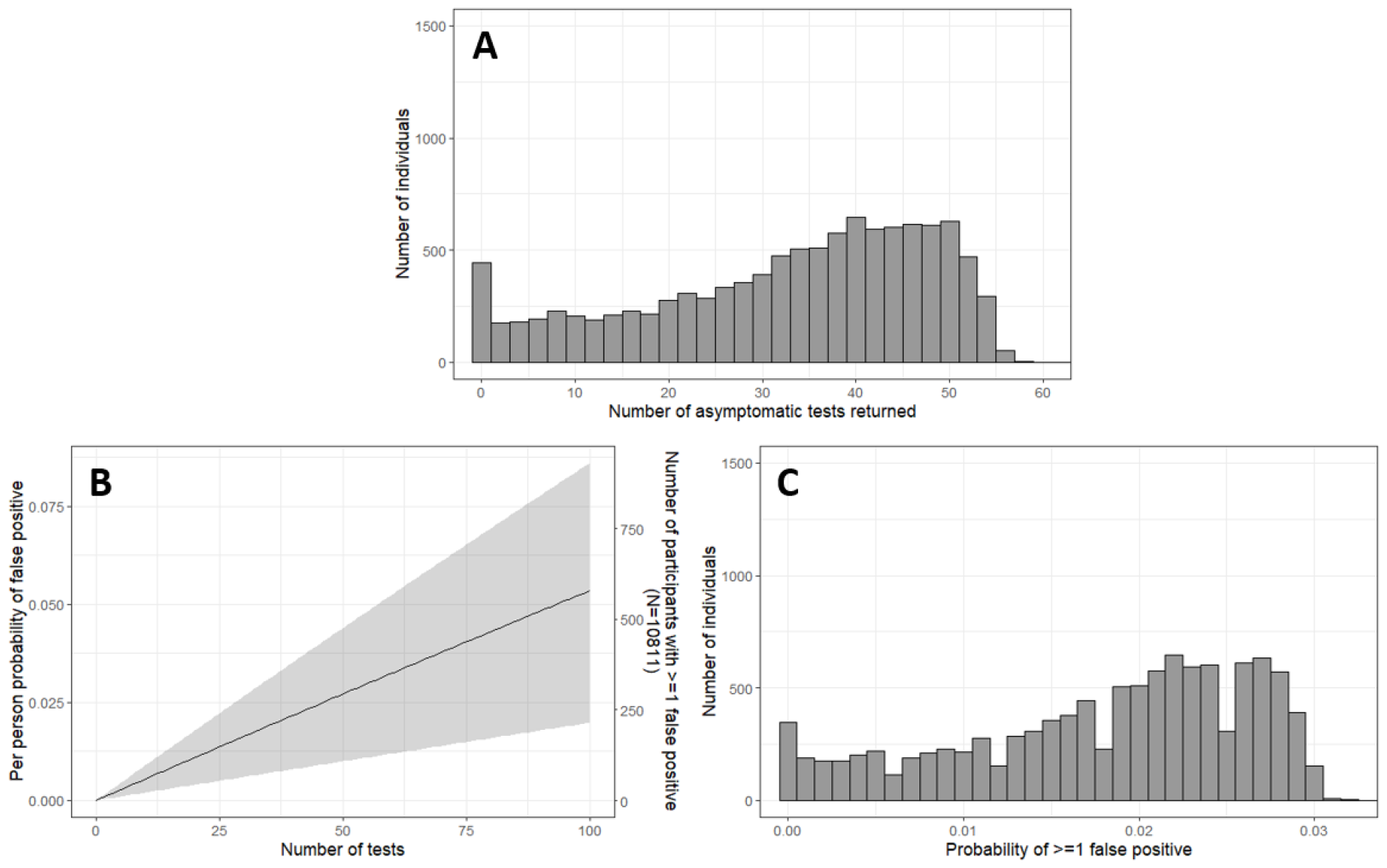
Estimated probability of false positives in all enrolled participants in the COV002 trial. A) Histogram of the number of asymptomatic tests returned by each participants, B) Assumed relationship between the number of asymptomatic tests returned and the probability of receiving at least one false positive result (line represents the estimated probability when test specificity is assumed to be 99·945%, and the upper and lower bounds of the grey band represent the estimates when specificity is 99·98% and 99·91%, respectively, C) Histogram of the participants’ estimated probability of receiving ≥ false positive, assuming 99·945% specificity.

## Discussion

Understanding the extent to which vaccines protect against infection and progression to more severe disease outcomes is important for understanding and predicting the epidemiological impact of vaccination programs. Regular testing of asymptomatic participants is logistically challenging and resource intensive, so is not usually used in phase III vaccine trials.^4^ In this paper, we described the implementation of regular asymptomatic testing in the first COVID-19 vaccine trial to use this approach and evaluated adherence to testing over time and across socio-demographic factors.

Adherence to asymptomatic testing in the COV002 trial was high, with a median of 75·0% of expected swabs returned over the 13-month swabbing period. This translated to a 74·5% probability of detecting an asymptomatic RT-PCR-positive specimen across the swabbing period and 78·8% over the period of VE evaluation. Adherence was highest over short term follow-up, and was maintained above 80% for 7 weeks after prime vaccination and over 60% for 33 weeks. This is consistent with other short-term asymptomatic COVID-19 testing schemes in university and hospital settings.^19, 20^ For example, in their study of weekly SARS-CoV-2 antigen testing in a university over a 6-week period, Bigouette *et al*. found weekly adherence ranged between 70·5% and 80·6%.^21^ Extremely high compliance was found in the Duke University asymptomatic SARS-CoV-2 infection surveillance testing programme, where 95% of weekly/twice-weekly tests were returned.^22^ In this study, students signed a pledge agreeing to rigorous testing and lost access to campus services if they missed scheduled tests. Our study adds to these by demonstrating the adherence that can be expected over a long follow-up, and where the reason for testing is to better understand a candidate vaccine as opposed to controlling transmission in an isolated population.

In the COV002 trial, adherence declined over time and after key events such as unblinding and infection. Automated weekly reminders were used to encourage adherence over time. Unlike university settings, restrictions such as cutting off access to facilities would not be possible or reasonable in the diverse population of a clinical trial. The odds of returning a swab were a further 31% lower after unblinding and 55% lower after testing positive. This demonstrates the well-known phenomenon that trial participant behaviour can differ in the absence of blinding or when perceived risk changes,^23^ underlining the importance of maintaining participant-blinding. Some participants were asked to stop testing after a positive swab by the UK contact tracing scheme, Test and Trace, which may have contributed to the observed reduction in testing. Our analysis raises questions on the value of asymptomatic follow-up over long time periods and following unblinding or infection, because if adherence becomes too low, the validity of VE estimates will be undermined.

Adherence also varied across socio-demographic characteristics. The swab return rate increased with age and decreased with BMI. It was higher in females, non-HCWs, participants of white ethnicity, and participants who were included in VE analysis. Lower adherence in males and younger people to COVID-19 preventative behaviours is well reported.^21, 24-27^ In their university serial testing study, Bigouette *et al*. found that the odds of returning at least 80% of weekly swabs increased by 16% per year of age, and that adherent students were more likely to be female (OR 2·07) and of white ethnicity (OR Black or Asian vs White = 0·6)^21^. This is important to consider when interpreting vaccine trial results, as participants with lower adherence will contribute less to VE estimates. If not adjusted for, this can bias the estimated VE if the characteristics that predict adherence are associated with protection.^14^ Participants with lower adherence may benefit from tailored strategies to maintain contribution during trials.

While we expect that a large proportion of true asymptomatic infections were detected, we also estimate that there were between 21 and 96 false positive asymptomatic infections during the period of VE evaluation. Considering in our VE analysis dataset that there were 285 asymptomatic infections, we do not expect the true number to be close to the top of this range. However, it is difficult to estimate the number of false positives using this strategy as it is highly sensitive to the assumed test specificity and other sources of contamination.^7, 14^ If the true value is close to our lower bound, the bias will be small. However, if approximately a third of the asymptomatic positives were false positives, the estimated VE would be significantly biased towards zero. Further research into the impact of incomplete detection of asymptomatic infections and false positives on VE estimates would be valuable.

The uncertainty in our estimation of false positives is one this paper’s limitations. Alternative approaches may be better, such as limiting samples to those below a threshold CT value or those positive in multiple gene targets.^28^ Our estimation of the change in odds of swab return over time across age groups uses reference characteristics rather than overall trial population characteristics. This allows for the individual effect of each variable to be demonstrated but gives a larger temporal change in odds than observed in the data. Additionally, as the trial was conducted in a unique context of a pandemic, our overall estimate for adherence may not be relevant in other contexts. Despite this, we believe our conclusions on differences adherence across socio-demographic characteristics, over time, and after infection or unblinding, are likely to be generalisable.

There were limitations associated with regular testing in the COV002 trial. Firstly, the infrastructure required would not have been logistically or financially feasible outside of a pandemic. Trial procedures undoubtedly placed considerable burden on participants and at times created anxiety regarding isolation procedures for those who exhibited prolonged viral shedding. As PCR test availability within the UK varied throughout the testing period, access to self-testing was often a benefit for participants but it may have interfered with participants presenting to the symptomatic pathway for the trial’s primary endpoint. It is also possible that participants shared their test kits with friends and family, though due to the way in which the tests were registered electronically using personal details we are confident that this was usually detected, and these tests excluded. It is unknown what impact weekly self-swabbing had on adherence to other trial procedures and whether it contributed to trial retention or withdrawal, though overall trial withdrawals were low during the testing period.

However, there are other benefits to this testing strategy. It was unknown whether the vaccine would affect viral transmission and if so, how this might be mediated. The data from weekly swabbing demonstrated that asymptomatic infection occurred in both controls and vaccinated participants, and that the viral dynamics in vaccinated participants suggested vaccination may reduce transmission, which was later demonstrated.^29^ Continued testing also allows for detection of waning VE and reduction of VE against new variants. These are important data to inform testing strategies, infection control practices and vaccination programmes.

In conclusion, the COV002 trial was the first COVID-19 vaccine trial to estimate VE for asymptomatic and overall disease through regular asymptomatic testing of trial participants. High adherence to testing suggests that trial participants were motivated over a prolonged period. Regular self-swabs performed at home can provide a higher level of surveillance compared with clinic visits and home-based practices could be considered for other trial procedures. Adherence did decline over time, highlighting the need for consideration regarding the optimal time over which intensive sampling provides valuable data. There was variation across socio-demographic groups and a greater understanding of the factors that contribute to adherence may help with future study design. While a large proportion of asymptomatic infections are expected to have been detected, we also estimate a substantial number of false positives. This information could be valuable for trial methodologists when designing future phase III vaccine trials.

## Data Availability

Anonymised participant data is available upon request directed to the alternative corresponding author. Proposals will be reviewed and approved by the sponsor, investigator, and collaborators on the basis of scientific merit. After approval of a proposal, data can be shared through a secure online platform after signing a data access agreement. All data will be made available for a minimum of 5 years from the end of the trial.

## Acknowledgements

The COV002 study was funded by UK Research and Innovation, National Institute for Health Research (NIHR), Coalition for Epidemic Preparedness Innovations, NIHR Oxford Biomedical Research Centre, Thames Valley and South Midland’s NIHR Clinical Research Network, and AstraZeneca. This report is independent research funded by the UK NIHR and UK Research and Innovation.

This work was supported by payments made to the Imperial College London, MRC Centre for Global Infectious Disease Analysis (reference MR/X020258/1), funded by the UK Medical Research Council (MRC). This UK funded award is carried out in the frame of the Global Health EDCTP3 Joint Undertaking. This work was also supported by the MRC Doctoral Training Partnership (to LRW and KRWE) and the Imperial College President’s Scholarship (to LRW).

We are grateful to the NIHR infrastructure provided through the NIHR Biomedical Research Centres and the NIHR Clinical Research Network at the UK study sites. We are grateful for the support of the Department of Health and Social care and the Office for Life Sciences for swabbing kits and access to PCR testing and the dedication of the Lighthouse laboratory teams. The views expressed in this publication are those of the author(s) and not necessarily those of the National Institute for Health Research or the Department of Health and Social Care. AstraZeneca reviewed the data from the study and the final manuscript prior to submission, but the authors retained editorial control. We thank the volunteers who participated in this study.

