## Supplementary materials for "Implementation and Adherence to Regular Asymptomatic Testing in a COVID-19 Vaccine Trial"

#### Supplementary methods

##### RT-PCR Test Processing

Tests were processed by Department of Health and Social Care (DHSC) Lighthouse laboratories, principally Glasgow, Alderley Park (Manchester) and Milton Keynes which all used the same RT-PCR assay described below. RT-PCR for three SARS-CoV-2 target genes (Nucleocapsid (N), Spike (S) and ORF1ab (ORF)) used the Thermo Fisher TaqPath RT-PCR COVID-19 assay (Thermo Fisher Scientific, Waltham, MA, USA), analysed using UgenTec Fast Finder 3.300.5 (TaqMan 2019-nCoV Assay Kit V2 UK NHS ABI 7500 v2.1, UgenTec, Cambridge, MA, USA). The Assay Plugin contains an Assay specific algorithm and decision mechanism that allows conversion of the qualitative amplification Assay PCR raw data from the ABI 7500 Fast into test results with minimal manual intervention. Samples were called positive in the presence of at least single N gene and/or ORF1ab but may be accompanied with S gene (1, 2 or 3 gene positives). S gene was not considered a reliable single gene positive (as of mid-May 2020).

##### Matching and Data Capture

A data agreement between NHS Digital and the University of Oxford allowed data matching between barcoded swabs and trial participants by the study data team. Positive weekly swab results were uploaded to the REDCap® trial database [1, 2]. This did not occur in real time. Participants received results directly from DHSC via email or text message. All participants received standardised weekly reminders by email or text message from their study site to remind them to return their home kits each week. The NHS swab dataset was then matched by code written by the trial statistics team in R statistical software (version 4.1.2, R Foundation for Statistical Computing, Vienna, Austria [3]) to the trial dataset. Matching was defined by at least one of six combinations whereby at least two personal identifiers matched (such as name, NHS number or date of birth).

##### Estimating probability of asymptomatic infection detection

We calculated individual-level probabilities of asymptomatic infection detection using COV002 swabbing data and data from Hellewell *et al.* (2021)[4] on PCR test sensitivity to asymptomatic SARS-CoV-2 infections. We used the data on PCR sensitivity to asymptomatic infections over the course of infection (Supplementary Table 1) to estimate the probability of asymptomatic infection detection for different frequencies of testing. We assumed an individual tested on a random day in the first week after infection, and on the same day of the week in future. Then for weekly testing with 100% adherence, the probability of asymptomatic infection detection was 83.3% (Supplementary Table 2).

To estimate the sensitivity of asymptomatic infection detection in each individual, we estimated the proportion of expected weekly asymptomatic tests they returned. We assumed that tests were taken on the same random day of the week for a given individual, as above. We then assumed that the weeks on which tests were returned were chosen randomly among the possible weeks, given the number of tests returned for the individual. The probability of asymptomatic infection detection is then the probability if tests were returned in all possible weeks, multiplied by the proportion of weeks in which tests were returned

#### Supplementary Tables

**Supplementary Table 1. PCR test sensitivity to asymptomatic infection detection, estimated in Hellewell et al.** [4]

| Days since infection | Median | Lower | Upper |
|---|---|---|---|
| **0** | 0.005 | 0.000 | 0.020 |
| **1** | 0.039 | 0.005 | 0.161 |
| **2** | 0.246 | 0.048 | 0.766 |
| **3** | 0.710 | 0.226 | 0.881 |
| **4** | 0.785 | 0.559 | 0.892 |
| **5** | 0.767 | 0.613 | 0.885 |
| **6** | 0.725 | 0.574 | 0.857 |
| **7** | 0.677 | 0.529 | 0.818 |
| **8** | 0.626 | 0.484 | 0.771 |
| **9** | 0.571 | 0.437 | 0.720 |
| **10** | 0.515 | 0.390 | 0.660 |
| **11** | 0.459 | 0.344 | 0.597 |
| **12** | 0.404 | 0.296 | 0.534 |
| **13** | 0.351 | 0.252 | 0.469 |
| **14** | 0.302 | 0.211 | 0.411 |
| **15** | 0.257 | 0.170 | 0.357 |
| **16** | 0.217 | 0.137 | 0.311 |
| **17** | 0.180 | 0.108 | 0.268 |
| **18** | 0.149 | 0.083 | 0.229 |
| **19** | 0.122 | 0.065 | 0.198 |
| **20** | 0.100 | 0.050 | 0.168 |
| **21** | 0.082 | 0.038 | 0.144 |
| **22** | 0.066 | 0.028 | 0.124 |
| **23** | 0.054 | 0.021 | 0.106 |
| **24** | 0.044 | 0.016 | 0.091 |
| **25** | 0.035 | 0.012 | 0.078 |
| **26** | 0.028 | 0.009 | 0.066 |
| **27** | 0.023 | 0.006 | 0.056 |
| **28** | 0.018 | 0.005 | 0.048 |
| **29** | 0.014 | 0.004 | 0.041 |
| **30** | 0.012 | 0.003 | 0.035 |

**Supplementary Table 2. Calculation of the probability of asymptomatic infection detection for weekly PCR testing.**

| First test (days since infection) | Equation | Median | Lower CI | Upper CI |
|---|---|---|---|---|
| **Day 0** | =1-(1-0.005)*(1-0.677)*(1-0.302)*(1-0.082)*(1-0.018) | 0.798 | 0.645 | 0.915 |
| **Day 1** | =1-(1-0.039)*(1-0.626)*(1-0.257)*(1-0.066)*(1-0.014) | 0.754 | 0.587 | 0.896 |
| **Day 2** | =1-(1-0.246)*(1-0.571)*(1-0.217)*(1-0.054)*(1-0.012) | 0.763 | 0.549 | 0.961 |
| **Day 3** | =1-(1-0.710)*(1-0.515)*(1-0.180)*(1-0.044) | 0.890 | 0.585 | 0.973 |
| **Day 4** | =1-(1-0.785)*(1-0.459)*(1-0.149)*(1-0.035) | 0.905 | 0.738 | 0.969 |
| **Day 5** | =1-(1-0.767)*(1-0.404)*(1-0.122)*(1-0.028) | 0.881 | 0.748 | 0.960 |
| **Day 6** | =1-(1-0.725)*(1-0.351)*(1-0.100)*(1-0.023) | 0.843 | 0.699 | 0.940 |
| **Average** | | 0.833 | 0.650 | 0.945 |

**Supplementary Table 3. Health and socio-demographic characteristics of all participants eligible for VE analysis in the COV002 phase III vaccine trial.**

| | VE dataset (N=8548) | VE dataset <75% swabs (N =2636) | VE dataset ≥75% swabs (N = 5912) |
|---|---|---|---|
| **Age, years** | | | |
| **<25** | 383 (4.5) | 234 (8.9) | 149 (2.5) |
| **25-34** | 1802 (21.1) | 822 (31.2) | 980 (16.6) |
| **35-44** | 2081 (24.3) | 759 (28.8) | 1322 (22.4) |
| **45-54** | 2186 (25.6) | 56.1 (21.3) | 1625 (27.5) |
| **55-64** | 910 (10.6) | 158 (6.0) | 752 (12.7) |
| **≥65** | 1186 (13.9) | 102 (3.9) | 1084 (18.3) |
| **Sex** | | | |
| **Male** | 3474 (40.6) | 1103 (41.8) | 2371 (40.1) |
| **Female** | 5074 (59.4) | 1533 (58.2) | 3541 (59.9) |
| **Ethnicity** | | | |
| **White** | 7898 (92.4) | 2328 (88.3) | 5570 (94.2) |
| **Asian** | 425 (5.0) | 197 (7.5) | 228 (3.9) |
| **Black** | 41 (0.5) | 15 (0.6) | 26 (0.4) |
| **Other** | 184 (2.2) | 96 (3.6) | 88 (1.5) |
| **BMI kg/m²** | | | |
| **Underweight** (<18.5) | 80 (0.9) | 17 (0.6) | 63 (1.1) |
| **Healthy weight** (18.5 to 24.9) | 3747 (43.8) | 1106 (42.0) | 2641 (44.7) |
| **Overweight** (25 to 29.9) | 3008 (35.2) | 901 (34.2) | 2107 (35.6) |
| **Obese** (≥30) | 1713 (20.0) | 612 (23.2) | 1101 (18.6) |
| **Occupation**[1] | | | |
| **Non-essential** | 1293 (15.1) | 166 (6.3) | 1127 (19.1) |
| **Essential, non-HCW** | 1642 (19.2) | 528 (20.0) | 1114 (18.8) |
| **HCW (0 COVID-19 patients)** | 3925 (45.9) | 1264 (48.0) | 2661 (45.0) |
| **HCW (1+ COVID-19 patients)** | 1688 (19.7) | 678 (25.7) | 1010 (17.1) |

Data are N (%). BMI = Body mass index, HCW = Healthcare worker. [1]Non-essential occupation classified as: Unemployed, Retired, Student, Hospitality/retail workers in non-essential shops and branches, Construction workers and labourers, or Other occupation, Essential, non-HCW occupation = Cleaning and domestic workers, Drivers and transport workers, Public safety workers (e.g. police, fire services, security), Religious workers, and Retail workers in essential shops and branches (e.g. food, chemists, banks).

**Supplementary Table 4. Number of positive, negative, and indeterminate tests returned in the full analysis dataset and the vaccine efficacy (VE) subset.**

| | Positive | Negative | Indeterminate | Total |
|---|---|---|---|---|
| **Full dataset** | 1636 | 348395 | 6520 | **356551** |
| **VE dataset** | 339 | 105101 | 2201 | **107641** |

**Supplementary Table 5. Multivariable mixed model results for adherence to weekly asymptomatic testing (including interactions with time).** Overall category P values calculated with type III ANOVA.

| | OR | LCI | UCI | P value |
|---|---|---|---|---|
| **Time since prime (weeks)** | 0.89 | 0.88 | 0.90 | <0.001 |
| **Age (vs <25 years)** | | | | <0.001 |
| 25-34 years | 1.64 | 1.24 | 2.17 | <0.001 |
| 35-44 years | 2.09 | 1.58 | 2.75 | 0.001 |
| 45-54 years | 2.25 | 1.7 | 2.97 | <0.001 |
| 55-64 years | 2.89 | 2.1 | 3.96 | <0.001 |
| 65+ years | 3.61 | 2.63 | 4.96 | <0.001 |
| **Occupation (vs non-HCW)** | | | | <0.001 |
| HCW, 0 COVID-19 patients | 0.67 | 0.59 | 0.76 | <0.001 |
| HCW, 1+ COVID-19 patients | 0.52 | 0.45 | 0.6 | <0.001 |
| **Male (vs female)** | 0.68 | 0.6 | 0.77 | <0.001 |
| **Non-white ethnicity (vs white)** | 0.44 | 0.37 | 0.52 | <0.001 |
| **BMI category (vs healthy weight)** | | | | <0.001 |
| Obese | 0.70 | 0.59 | 0.82 | <0.001 |
| Overweight | 0.86 | 0.75 | 0.98 | 0.028 |
| Underweight | 0.94 | 0.51 | 1.72 | 0.843 |
| **Comorbidity (yes)** | 1.32 | 1.14 | 1.53 | <0.001 |
| **Included in VE analysis (no)** | 0.29 | 0.25 | 0.34 | <0.001 |
| **Other swab (yes)** | 1.31 | 1.13 | 1.52 | <0.001 |
| **Follow-up period (vs during follow-up)** | | | | <0.001 |
| Pre-VE follow up | 0.53 | 0.51 | 0.56 | <0.001 |
| Post-unblinding | 0.65 | 0.63 | 0.68 | <0.001 |
| Post-positive swab | 0.46 | 0.41 | 0.52 | <0.001 |
| Post-December 8th | 0.93 | 0.89 | 0.98 | 0.003 |
| **Age (vs <25 years) x Time since prime (weeks)** | | | | <0.001 |
| 25-34 years | 1.02 | 1.01 | 1.02 | <0.001 |
| 35-44 years | 1.03 | 1.03 | 1.04 | <0.001 |
| 45-54 years | 1.05 | 1.04 | 1.06 | <0.001 |
| 55-64 years | 1.06 | 1.05 | 1.07 | <0.001 |
| 65+ years | 1.08 | 1.07 | 1.09 | <0.001 |
| **Male (vs female) x Time since prime (weeks)** | 1.00 | 1.00 | 1.01 | 0.004 |
| **BMI category (vs healthy weight) x Time since prime (weeks)** | | | | <0.001 |
| Obese | 0.99 | 0.99 | 0.99 | <0.001 |
| Overweight | 1.00 | 0.99 | 1.00 | 0.038 |
| Underweight | 1.02 | 1.01 | 1.04 | 0.001 |
| **Comorbidity (yes) x Time since prime (weeks)** | 0.99 | 0.99 | 1.00 | 0.001 |
| **Included in VE analysis (no) x Time since prime (weeks)** | 0.99 | 0.98 | 0.99 | <0.001 |

### Supplementary figures

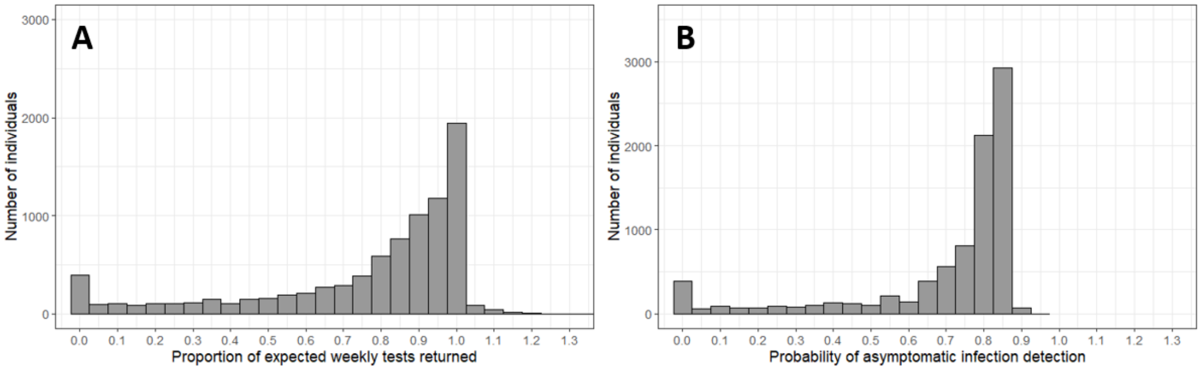

**Supplementary Figure 1. Adherence and estimated probability of asymptomatic infection detection in all participants in the COV002 trial eligible for VE analysis, during the period of VE follow up.** A) Histogram of participants' adherence to weekly testing, B) Histogram of the participants' estimated probability of asymptomatic infection detection. Probabilities over one show participants who returned more than the requested one swab per week.

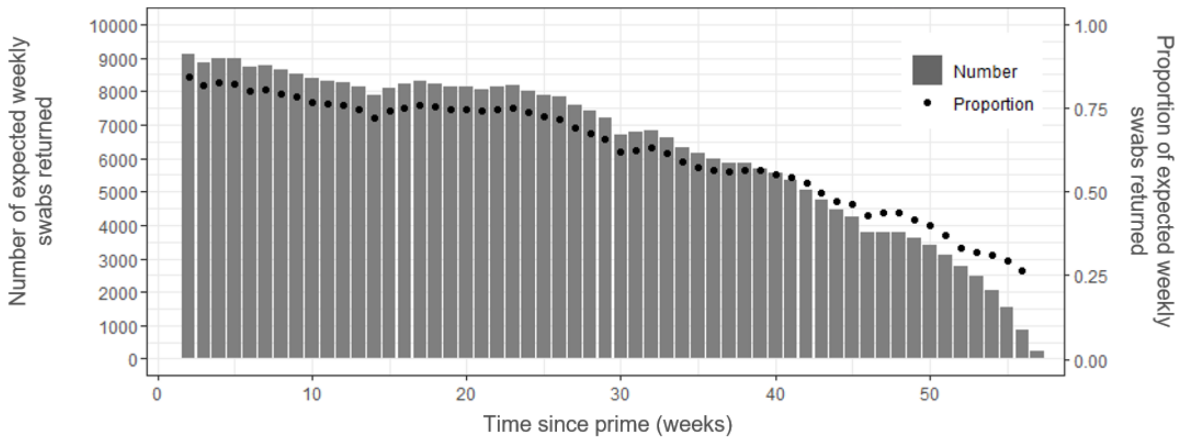

**Supplementary Figure 2. Number and proportion of swabs returned over time since prime vaccination.**

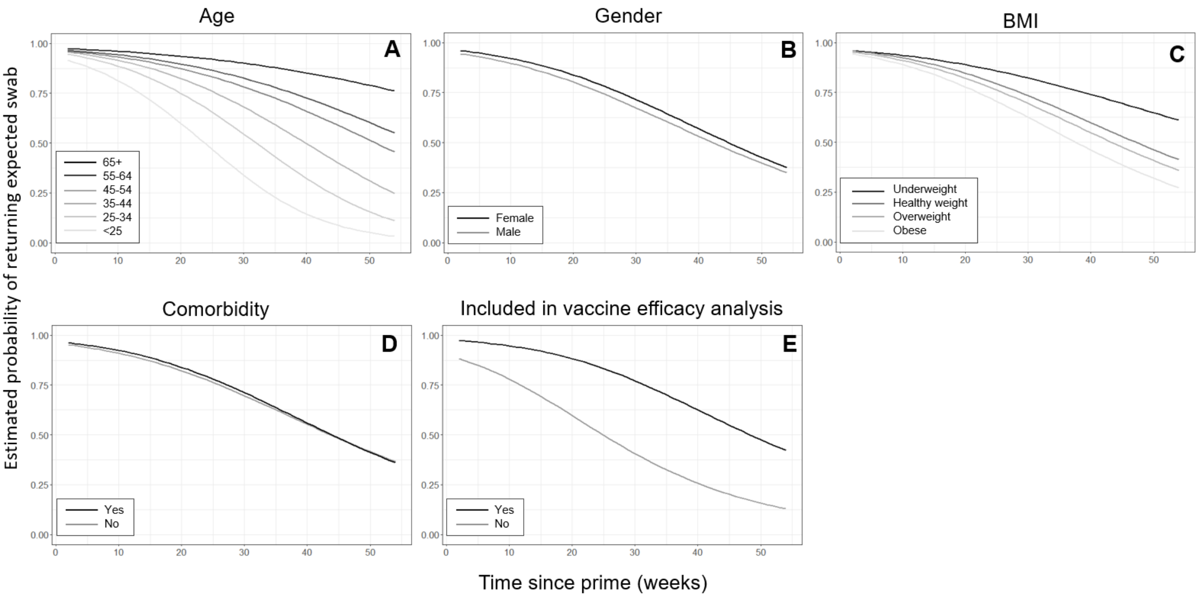

**Supplementary Figure 3. Model predicted probability of returning an expected asymptomatic test over time, by different socio-demographic variables.** A) age, B) gender, C) BMI, D) Comorbidity, E) Inclusion in vaccine efficacy (VE) analysis. Participants were included in VE analysis if they were enrolled in the efficacy cohort, were seronegative at baseline and did not test positive before the start of VE follow-up. Predictions made for the true COV002 population characteristics. One variable was set constant for each graph to show the effect of all participants having the characteristic of interest. Time dependent variables set constant: pre-VE follow up = no, post-unblinding = no, post-positive swab = no, post-December 8$^{th}$ = no.

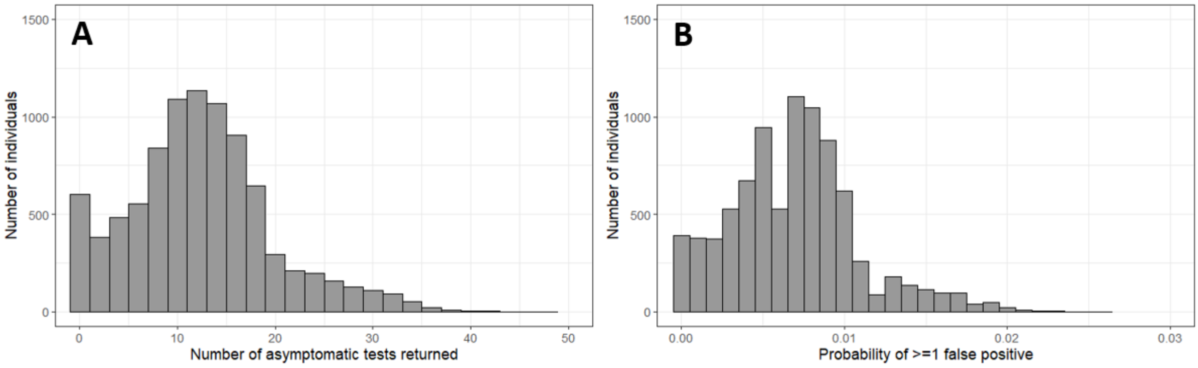

**Supplementary Figure 4. Adherence and estimated probability of false positives in all participants in the COV002 trial eligible for VE analysis, during the period of VE follow up.** A) Histogram of the number of asymptomatic tests returned by each participant, B) Histogram of the participants' estimated probability of receiving ≥ false positive.
